# Abnormal sleep blood pressure patterns are associated with diffusion tensor imaging along perivascular spaces index in cognitively impaired individuals

**DOI:** 10.1101/2025.02.23.25322732

**Authors:** Mariateresa Buongiorno, Gonzalo Sánchez-Benavides, Giovanni Caruana, Andrea Elias-Mas, Cristina Artero, Natalia Cullell, Pilar Delgado, Darly Milena Giraldo, Clara Marzal-Espí, Oriol Grau-Rivera, Alejandro de la Sierra, Ariane Delgado-Sanchez, Nicola J. Ray, Jerzy Krupinski

## Abstract

Blood pressure (BP) physiologically dips during sleep, and lack of dipping associates with adverse health outcomes and cognitive decline. Vascular pulsatility is the main driver of glymphatic cerebrospinal fluid transport, which removes metabolic waste products from the brain during sleep. We hypothesized that abnormal sleep BP patterns may affect glymphatic system health, and that this relationship may result in lower diffusion tensor imaging along perivascular spaces (DTI-ALPS) indices, a proposed neuroimaging index of glymphatic health. Twenty-one participants with mild-to-moderate cognitive impairment underwent 24-hour ambulatory BP monitoring, DTI-MRI and Alzheimer’s disease (AD) biomarker studies. Eight participants were classified as dippers (≥10%) and 13 as non-dippers (<10%), using sleep/awake systolic BP percentage of change. We found that non-dippers had lower DTI-ALPS even when adjusted by age and clinical stage (p=0.013). Stiffness measures (pulse wave velocity) were negatively correlated to DTI-ALPS (r=-0.5), but the association disappeared when adjusted by age. Positive AD biomarkers were more frequent in individuals who were classified as non-dippers of both systolic and diastolic BP, as compared to systolic and diastolic dippers (p=0.041). Our findings suggest that deviations of the physiological dipping sleep BP pattern may relate to poorer glymphatic function and increased AD pathology.

## Introduction

The circadian pattern of blood pressure (BP), measured through 24-hour ambulatory BP monitoring (ABPM), typically shows higher values during the day and a 10% to 20% reduction during sleep, a phenomenon known as “dipping”.^1,2^ Absence of this normal dipping pattern -non-dipping-, has been associated with future adverse health outcomes, including increased cardiovascular risk^3,4^ and cognitive decline.^5^ In a recent systematic meta-analysis, Gavriilaki and colleagues found that individuals with a normal dipping pattern had a 63% lower risk of all-cause dementia compared to non-dippers.^5^ Moreover, those classified as “risers” (non-dippers with even higher nighttime than daytime BP) were found to have a sixfold higher risk of abnormal cognitive function compared to dippers. Notably, a longitudinal study over 24 years in more than 1500 Swedish older men, found that the riser pattern was linked to increased likelihood of being diagnosed with any dementia and Alzheimer’s disease (AD) but not vascular dementia.^6^ Such findings suggest the existence of mechanisms that may act as risk factors for AD independently from direct cerebrovascular damage.

Studies focused on patients already diagnosed with AD revealed that abnormal ABPM patterns (non-dipper, riser, or extreme dipper) are highly prevalent (>80%),^7,8^ compared to matched controls (38%).^7^ Similarly, individuals with mild cognitive impairment (MCI), the transitional stage between intact cognition and dementia, seem to exhibit higher nighttime systolic BP compared to controls,^9^ and presence of MCI is more frequent in extreme dippers (32.0%), non-dippers (30.0%) and risers (50.0%) than in dippers (13.2%).^10^

Research exploring the association between ABPM patterns and pathophysiological markers of AD is very limited. Tarumi and colleagues reported that among amnestic MCI patients, those with a non-dipping pattern showed increased levels of amyloid-β (Aβ) deposition in the posterior cingulate, a key region in early accumulation of pathology in AD, as measured by 18F-florbetapir PET SUVR.^11^

One hypothesis for the association between abnormal sleep BP and cognitive decline revolves around the potential impact on the glymphatic system, a brain-wide waste clearance pathway that is primarily active during sleep. The glymphatic system relies heavily on vascular pulsatility, generated by the rhythmic contractions of arteries, to drive cerebrospinal fluid (CSF) through perivascular spaces, facilitating the removal of metabolic waste products from the brain.^12,13^ Mestre et al. demonstrated in 2018 that CSF flow was pulsatile and synchronized with the cardiac cycle using particle tracking in live mice, with arterial wall motion, driven by perivascular pumping, as the primary mechanism.^14^ Moreover, increased BP altered arterial wall pulsations, increasing backflow-in which CSF travels back towards arterial spaces thereby reducing net CSF flow.^14^ As cerebral arterial pulsatility is a major driver of CSF influx through the brain parenchyma and sleep plays a critical role, we hypothesize that deviations from normal blood pressure pattern during sleep may plausibly impair the efficiency of waste clearance via the glymphatic pathway.

To our knowledge, no direct evidence linking sleep BP patterns with glymphatic function has been established. This study aimed to investigate the relationship between dipping and non-dipping sleep BP patterns and diffusion tensor imaging along perivascular spaces (DTI-ALPS), a postulated neuroimaging proxy for glymphatic function, in individuals with cognitive impairment. Our main hypothesis was that any deviation from sleep dipping BP pattern will be associated with a lower DTI-ALPS index, suggesting poorer glymphatic functioning.

## Methods

### Participants

The sample was composed of 21 participants with cognitive impairment (either amnestic MCI or dementia) with at least 0.5 in Clinical Dementia Rating (CDR), recruited between December of 2022 and December 2023 from the Cognition and Behavior Unit at the Department of Neurology, Hospital Universitari MútuaTerrassa, Barcelona, Spain. More details have been provided in Ray et al., 2024. ^15^

### Clinical measures

Participants were staged using global CDR score. Global cognitive performance was assessed using the mini-mental state examination (MMSE), and episodic memory with the Repeatable Battery for the Assessment of Neuropsychological Status (RBANS) delayed memory index adjusted by age (reference population mean=100, SD=15).

### Ambulatory BP data collection and definition of BP night patterns

ABPM was performed for 24 hours using the BP monitor Mobil-O-Graph® PWA (I.E.M. Industrielle Entwicklung Medizintechnik GmbH, Stolberg, Germany). Data were processed with the provided Hypertension Management Software Client-Server version 5.2.2 (HMS CS 5.2.2). The procedure was as follows: between 9:00AM and 11:00AM the participants came to the clinic and the device was placed in the non-dominant arm. They were instructed to avoid efforts during BP takings and recording any incidence in a provided diary. They came back the morning after at the same time approximately. The device was removed, and the data downloaded. Participants were instructed to record sleep and wake time in the diary, as well as any incidence during takings, with the assistance of their relatives if necessary. BP measures were taken every 20 minutes for 24h. Only valid measures were used in the computation of awake and sleep periods mean BP. We averaged BP measures considering the actual sleep/awake time reported in the diary, and calculated the relative magnitude of BP changes from awake to sleep period using the formula:

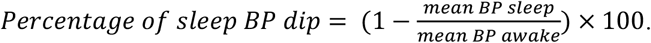

We categorized systolic BP (SBP) sleep patterns following standard references in two main categories: dipping (sleep SBP dip ≥10%) and non-dipping (sleep SBP dip <10%). Extreme patterns were also labelled within each group: extreme dipping (sleep SBP dip >20%) and rising (sleep SBP dip <0%).

Since we hypothesized that any deviation of the physiological patterns of sleep BP may be deleterious for glymphatic function, we also defined groups for diastolic BP (DBP) and, for secondary analyses, we created a Normal sleep BP pattern when both SBP and DBP showed a “dipping” pattern and considered an abnormal sleep BP pattern any deviation from “dipping”. Two groups within the abnormal patterns were created: One Abnormal was defined when only one abnormal pattern (either SBP or DBP different from dipping) was found, and Two Abnormal when both SBP and DBP sleep pattern were different from dipping.

Peripheral stiffness was explored using the mean of the augmentation index (AIx) corrected for a heart rate of 75 bpm (AIx@75). Central stiffness was measured through and pulse wave velocity (PWV).

### MRI acquisition and DTI-ALPS calculation

Scans were acquired using a 3T MR scanner (Philips Ingenia Elition). A standardized MR protocol was used for the acquisition, including a diffusion weighted imaging (DWI) sequence (TR = 2.53, TE = 0.07, slice thickness = 2.2 mm, voxel size= 1.69 × 1.69 × 2.2. DWI was performed in 128 directions (diffusion b=1000 s/mm2) and in one acquisition without diffusion weighting (B0). The DWI images were processed with FSL.^16^ Images were skull stripped^17^ and eddy current-induced distortions and subject movements were corrected.^18^ To calculate the ALPS-index FSL’s DTIFIT was used to create diffusivity maps in subject space in the x, y, and z directions, as well as a color-coded vector image showing the principal diffusion tensor direction (V1). To identify the location of the PVS, SWI were spatially co-registered with the subject-space B0 image using ANTs and superimposed on the V1 image to identify the medullary veins located perpendicularly to the PVSs and within the projection and association fibers. Two 4 mm spheres were placed at these locations, and diffusion in x, y and z directions spheres were extracted and used to compute the ALPS index^19^ according to the following equation:

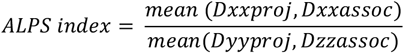

### AD biomarker status

Presence of AD pathology was defined as having a positive visual read in amyloid PET (^18^F-Flutemetamol) as rated by an experienced nuclear medicine physician following standard guidelines. In one participant with PET data not available we used the CSF ratio t-tau/Aβ42, quantified with Lumipulse essay kits from Fujirebio (Fujirebio Inc. Europe, Gent, Belgium) using local cut-off values.^20^

### Statistical analyses

We compared demographic, clinical, AMBP variables, and DTI-ALPS index scores between dipping and non-dipping groups using Mann-Whitney’s tests for continuous variables and Fisher’s exact test for categorical ones. To further explore the group differences in DTI-ALPS we performed a rank-based regression accounting by the effect of age and CDR using the Rfit package. We also explored the association between DTI-ALPS and continuous sleep SBP changes through correlations.

In secondary analyses, we created three groups to better account for any deviation on the physiological sleep BP dipping in both SBP and DBP. We defined as abnormal any other pattern (i.e. non-dipping, riser, extreme dipping) and explored the differences among Normal (dipping pattern in both SBP and DBP), One Abnormal BP (abnormal pattern either in SBP or DBP), and Two Abnormal BP (abnormal pattern in both SBP and DBP), through Kruskal-Wallis and with further adjustments by age. We also assessed the association between sleep, awake and total Aix@75 and PWV measures, as proxies of vascular stiffness, and DTI-ALPS, using correlations.

A threshold of p<0.05 was used for statistical significance. P values <0.1 were considered as trends. Given the exploratory nature of the study, no adjustments by multiple comparisons were considered. Analyses and plots were performed using R statistical software.

## Results

### Prevalence of SBP dipping/non-dipping patterns and group differences

Thirteen participants (62%) out of the 21 recruited displayed a non-dipping sleep SBP pattern. Descriptive data for demographic, clinical, AMBP, and DTI-ALPS measures by dipping/non-dipping patterns are shown in Table 1. No differences in age, sex, CDR, and cognitive outcomes were found between SBP dipping and non-dipping groups, although non-dippers tend to show lower cognitive scores (MMSE, p=0.1) and a higher proportion of positive AD biomarkers (85% vs 50%; p=0.14). Dippers had higher level of education (p=0.04), lower mean sleep SBP measures (p=0.02), lower sleep PWV values (p=0.03).

**Table 1.** Demographic, clinical, cognitive, BP, and DTI-ALPS descriptives by sleep SBP patterns.

|  | SBP Dipping | SBP Non-dipping | *P* |
| --- | --- | --- | --- |
| Number of individuals | 8 | 13 |  |
| Age, years Median [IQR] | 72.5 [10.5] | 74.0 [4.0] | 0.44 |
| Sex, females n (%) | 5 (62.5%) | 10 (76.9%) | 0.63 |
| Education years Median [IQR] | 11.5 [5.0] | 7.0 [5.0] | **0.04** |
| MMSE Median [IQR] | 28.0 [6.5] | 21.0 [7.0] | 0.10 |
| RBANS delayed memory score Median [IQR] | 75 [28.5] | 54 [27] | 0.24 |
| CDR Global score |  |  | 0.71 |
| CDR 0.5 n (%) | 6 (75%) | 11 (84.6%) |  |
| CDR 1 n (%) | 1 (12.5%) | 2 (15.4%) |  |
| CDR 2 n (%) | 1 (12.5%) | 0 (0%) |  |
| AD biomarker status, positive n (%) | 4 (50%) | 11 (84.6%) | 0.14 |
| Hypertension, presence n (%) | 3 (37.5%) | 9 (69.2%) | 0.20 |
| Mean SBP awake Median [IQR] | 131.7 [17.3] | 124.8 [18.0] | 0.71 |
| Mean SBP sleep Median [IQR] | 104.9 [20.1] | 123.3 [18.5] | **0.02** |
| SBP sleep percent change Median [IQR] | 14.9 [5.6] | 1.7 [7.4] | **<0.001** |
| Mean DBP awake Median [IQR] | 74.8 [8.7] | 70.1 [14.5] | 0.24 |
| Mean DBP sleep Median [IQR] | 59.4 [5.2] | 63.2 [9.4] | 0.19 |
| DBP sleep percent change Median [IQR] | 22.5 [10.3] | 8.4 [8.5] | **0.003** |
| Aix@75 awake Median [IQR] | 25.5 [8.5] | 25.0 [10.0] | 0.66 |
| Aix@75 sleep Median [IQR] | 25.5 [7.5] | 36.0 [10.0] | 0.13 |
| Aix@75 total Median [IQR] | 26.0 [5.3] | 31.0 [11.0] | 0.26 |
| PWV awake Median [IQR] | 10.9 [2.1] | 10.9 [0.5] | 0.88 |
| PWV sleep Median [IQR] | 10.3 [1.7] | 10.9 [0.5] | **0.03** |
| PWV total Median [IQR] | 10.6 [1.8] | 11 [0.4] | 0.36 |
| DTI-ALPS index | 1.322 [0.186] | 1.146 [0.105] | **0.03** |

Significance was tested using Mann-Whitney test for continuous variables and Fisher's exact test for categorical variables. SBP: systolic blood pressure; IQR: interquartile range; MMSE: Mini-Mental State Examination; RBANS: repeatable battery for the assessment of neuropsychological status; CDR: clinical dementia rating; AD: Alzheimer's disease; DBP: diastolic blood pressure; Aix@75: augmentation index corrected for a heart rate of 75 bpm; PWV: pulse wave velocity; DTI-ALPS: diffusion tensor imaging along perivascular spaces

### DTI-ALPS differences between SBP dipping and non-dipping group

There were significant differences between dipping and non-dipping group in DTI-ALPS indices using a Mann-Whitney test (p=0.03). Such differences persisted after adjustment by age and CDR in rank-based regression (p=0.013). See Figure 1.

**Figure 1.**
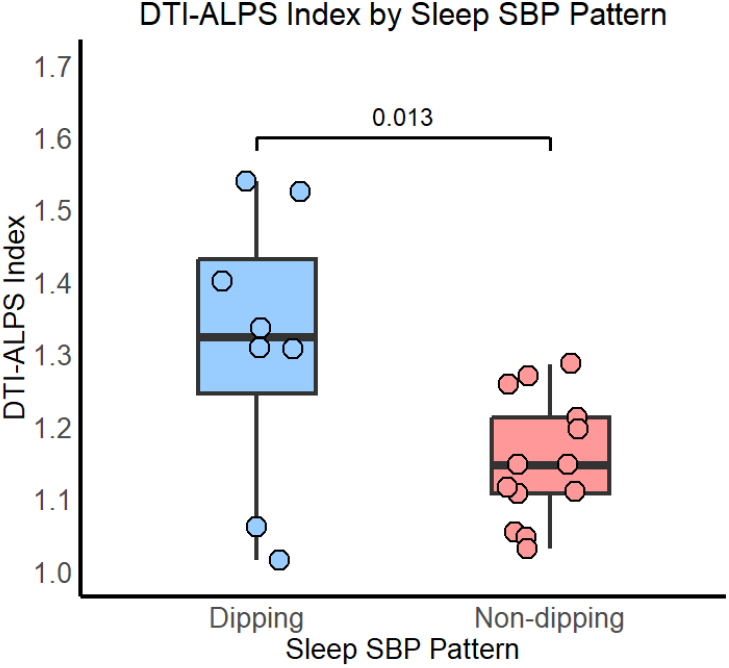
DTI-ALPS index by sleep SBP pattern. P-value adjusted by age and CDR.

### Associations between continuous sleep BP percentage of change and DTI-ALPS

Correlational analyses in the full sample revealed low positive associations between DTI-ALPS and the percentage of SBP change between awake and sleep that did not reach statistical significance (r=0.36, p=0.11). After removing extreme dippers (n=19), the associations were moderate and significant (r=0.49, p=0.03). See Figure 2. For DBP the results were similar, (full sample r=0.34, p=0.12; restricted sample n=16, r=0.57, p=0.024).

**Figure 2.**
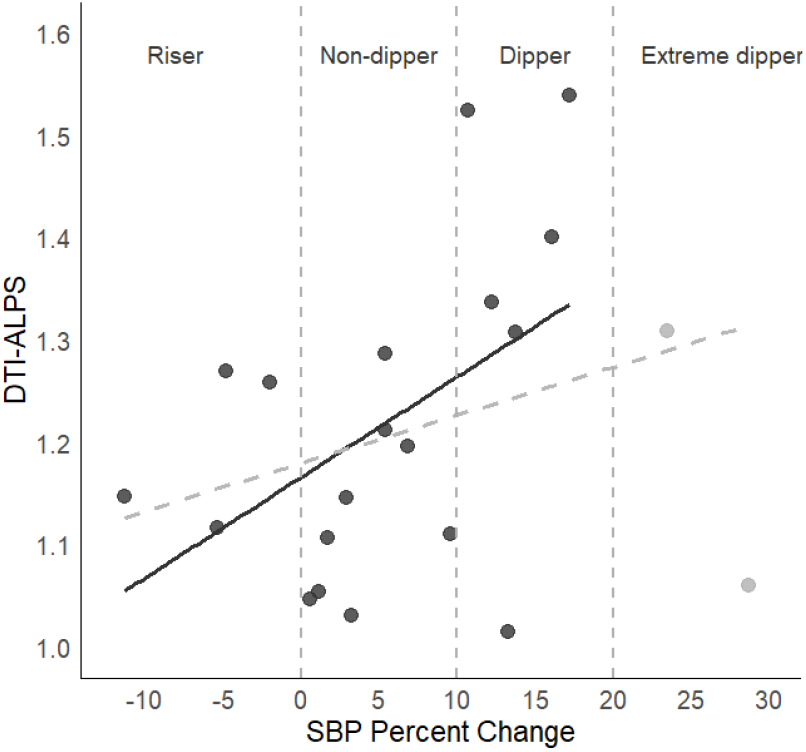
DTI-ALPS index and sleep SBP percentage of change. Dashed grey line shows the association in the full sample and black continuous line the association after removing the two individuals labelled as extreme dippers.

### Secondary analyses with normal and abnormal SBP/DBP sleep pattern groups

Four participants (19%) out of the 21 recruited displayed both SBP and DBP normal (dipping) sleep BP patterns. The remaining 17 participants (81%) had abnormal patterns, being the group with both SBP and DBP altered patterns the most prevalent (10 participants out of 21, 47.6%). The three groups (Normal, One Abnormal, and Two Abnormal) differed in the percentage of participants with positive AD pathophysiological biomarkers (p=0.043) and in DTI-ALPS index (p=0.024). Pairwise comparisons showed that the difference in percentage of AD positive biomarkers was driven by the Two Abnormal group as compared to the Normal group (p=0.041, One Abnormal vs Normal p=0.24). Similarly, DTI-ALPS index was significantly lower in the group with Two Abnormal BP patterns as compared to the Normal (p=0.004) and showed a trend when One Abnormal group was compared to the Normal one (p=0.073), see Figure 3. The significant difference between Two Abnormal BP patterns and Normal persisted after adjusting by age (p=0.005).

**Figure 3.**
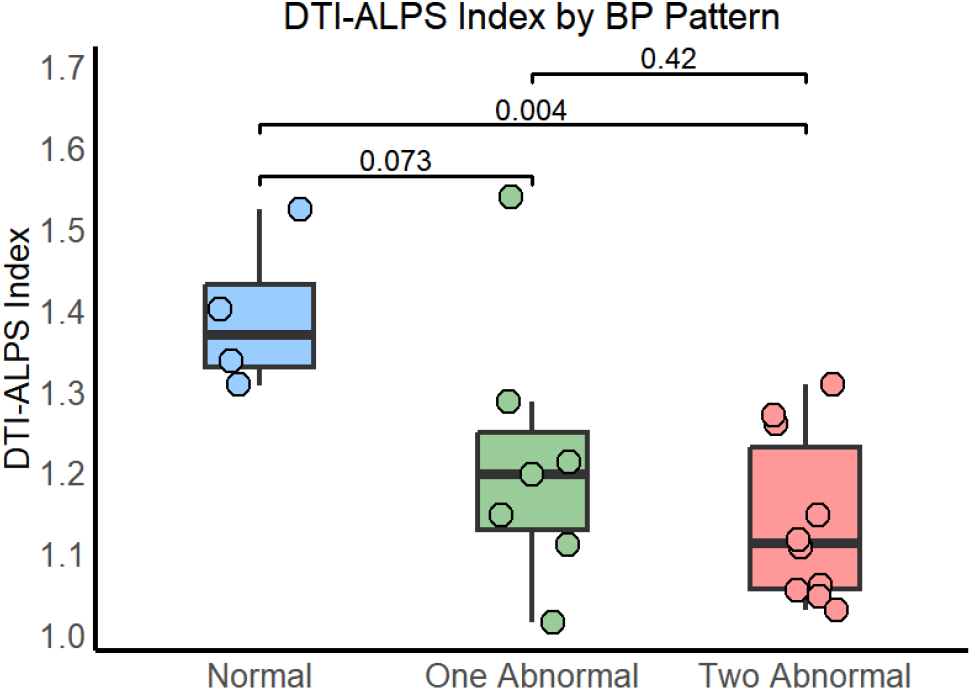
DTI-ALPS index by sleep BP pattern. Normal: dipping pattern in both SBP and DBP; One Abnormal: abnormal pattern either in SBP or DBP; Two Abnormal: abnormal pattern in both SBP and DBP.

### Associations between stiffness measures, BP groups, and DTI-ALPS

We observed higher PWV values during sleep in non-dipping than in dipping (p=0.03), that moderated when adjusted by age in rank-based regression (p=0.08). Correlational analyses between DTI-ALPS and PWV showed significant moderate negative correlations during awake and sleep measures (around r=-0.5, see Figure 4), but after adjusting by age, such associations disappeared (p>0.4). Regarding Aix@75, no significant associations were observed.

**Figure 4.**
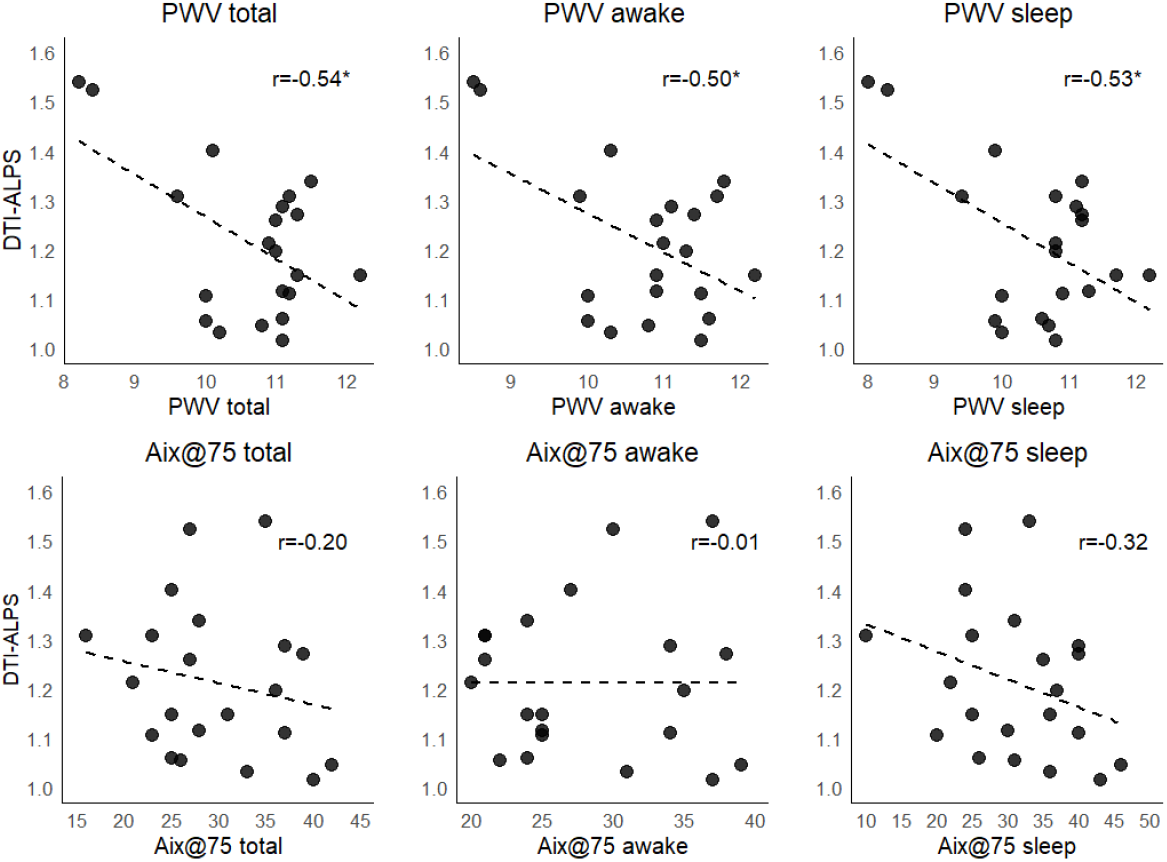
DTI-ALPS index unadjusted association with stiffness measures. PWV: pulse wave velocity; Aix@75: augmentation index corrected for a heart rate of 75 bpm

## Discussion

In this study, we investigated the associations between sleep BP patterns and glymphatic function, measured by the DTI-ALPS index, in individuals with cognitive impairment. Consistent with our hypothesis, the findings suggest that deviations from the physiological dipping BP pattern during sleep are associated with reduced glymphatic function, as reflected by lower DTI-ALPS indices.

The main result of this study showed that SBP non-dipping patterns were associated to lower DTI-ALPS indices. Results persisted after adjusting by age and clinical stage (CDR). When any deviation of the physiological dipping pattern was considered (either in SBP or DBP) we also found higher presence of AD pathology as measured with in-vivo biomarkers in individuals with abnormal patterns. Globally, our findings are in line with the body of literature supporting the relevance of vascular dynamics in glymphatic function. The glymphatic system relies on CSF transport through perivascular spaces, which is driven by arterial pulsatility, respiration, and slow vasomotion.^21^ Altered vascular dynamics, such as reduced arterial compliance and increased peripheral resistance, may contribute to impaired glymphatic clearance, thereby increasing the accumulation of neurotoxic proteins like Aβ and tau. Evidence in animal models have shown that CSF flow in perivascular spaces is primarily driven by arterial pulsations linked to the cardiac cycle, and that under conditions of elevated blood pressure, the efficiency of this arterial-driven CSF flow is reduced.^14^ Similarly, Mortensen et al. demonstrated that in spontaneous hypertensive rats, glymphatic transport is compromised, and solute clearance from brain parenchyma is decreased, being this effect worse in states of chronic hypertension.^22^

It is well stablished that glymphatic transport occurs mainly during sleep,^23^ and it has been also well documented that BP non-dipping patterns are associated to increased risk of cognitive decline and dementia.^5,6^ Our hypothesis suggests that the negative outcomes observed in presence of abnormal sleep BP patterns would act through glymphatic alterations and impaired solute clearance. Fultz et al. showed that during non-rapid eye movement sleep in healthy young humans, large oscillations in CSF flow are tightly coupled with neural and hemodynamic rhythms.^24^ Specifically, CSF flow oscillations are anticorrelated to a hemodynamic oscillation in the cortical gray matter that appears during sleep, with CSF flow increasing when blood volume decreases. Our findings of reduced DTI-ALPS indices in cognitively impaired older individuals with abnormal sleep BP patterns can be interpreted in the light of such observations: the observed altered hemodynamic oscillation at nighttime would in turn alter CSF flow oscillations, impairing glymphatic function as measured by DTI-ALPS. Thus, our data support for the first time the concept that the cognitive decline associated to the absence of physiological blood pressure drop during sleep would act through glymphatic dysfunction.

The association that we observed between the percentage of BP dip during sleep and DTI-ALPS, further support the concept that a greater nocturnal BP dip is linked to better glymphatic function. However, this association appears to be true within a range of BP change since the correlations strengthened and became significant (from r=0.3 to r=0.5) after excluding individuals with an extreme-dipping pattern (>20%). Extreme dipping has been also related to presence of cognitive impairment.^10^ This evidence, together with our findings, support the hypothesis that any deviation from the physiological sleep BP dipping range may adversely affect glymphatic function. Moreover, in line with the broader hypothesis suggesting that any alteration of the sleep vascular fitness may impair glymphatic function, we observed a negative association between stiffness measures (PWV) and DTI-ALPS. Although the associations disappeared when adjusted by age, our observations align with the reported link between PWV and increased risk of dementia,^25,26^ and higher amyloid deposition.^27,28^ Our results would also mirror recent evidence showing that PWV relates to enlarged perivascular spaces (ePVS).^29^ ePVS measures have been proposed as another neuroimaging proxy to glymphatic functioning, since PVS enlargement may occur due to CSF stagnation caused by the disruption of CSF flow.

Although no differences in the frequency of presence of positive AD biomarkers was found when we compared dippers and non-dippers based on SBP, we observed a higher percentage of individuals with altered AD biomarkers in participants with abnormal BP patterns in both SBP and DBP when any deviation of dipping pattern was used. This finding aligns with the hypothesis that abnormal BP patterns would impair protein clearance, increasing Aβ pathology, and mirror the findings reported by Tarumi and colleagues with amyloid PET data in MCI patients focused in SBP measures.^11^ To our knowledge, no other studies have analyzed the relationship between sleep BP patterns and AD pathological markers. Further studies are needed to elucidate the underpinnings of such associations. A study exploring other neuroimaging outcomes demonstrated that increased 24-h AMBP pulse-pressure was associated with DTI metrics, such as decreased fractional anisotropy and increased mean diffusivity, suggesting a deterioration in brain neuronal fiber integrity.^30^ However, in that study no association between nocturnal BP dip and DTI metrics were found. A more recent MRI study performed in a large Korean cohort of almost 1400 healthy individuals found that increased systolic or diastolic BP variability during night, but not mean BP, was associated with a reduction of temporal grey matter volume and cognitive decline after 4 years, but they did not specifically assess BP dipping patterns.^31^

Night BP pattern alterations may not be only produced by hypertension or vascular risk factors. The early neurodegeneration of locus coeruleus (LC) in AD and some alpha-synucleinopathies may also generate sleep BP abnormal patterns. LC, in addition to its known regulation of sleep-wake cycle, also innervates brain precapillary and capillary vessels, regulates neurovascular coupling, and controls astrocytes, endothelial cells and pericytes, which together constitute the neurovascular unit, core in the functioning of glymphatic system.^32^ Indeed, the magnitude of circadian pattern of BP, but not global BP, has been shown to be reduced after LC lesions in rats.^33,34^ The fact that, in AD, LC starts accumulating p-Tau decades before the onset of cognitive impairment,^35^ and that in Parkinson disease, LC disintegration has been related to dysautonomia^36^ (being dysautonomia associated with faster disease progression)^37^ suggests a relevant role of the link between LC and circadian BP in the etiology and progression of neurodegenerative diseases. In a previous work, we presented a model linking glymphatic dysfunction to the pathogenesis of alpha-synucleinopathies, emphasizing the role that vascular regulation disturbances may play in the heterogeneous progression of such diseases.^38^ The current findings further support this model by suggesting that abnormal sleep BP patterns significantly reduce glymphatic efficiency, as measured by DTI-ALPS. Our model proposed that glymphatic dysfunction, synergistically enhanced by sleep disturbances and vascular dysregulation, fosters a vicious cycle of protein accumulation and neurodegeneration. The observed association between abnormal BP patterns and reduced DTI-ALPS indices in the present study strengthens the argument that vascular dynamics, regulated by LC, is crucial for maintaining glymphatic efficiency. Importantly, this proposal expands the etiology of BP abnormalities beyond purely vascular risk factors (i.e., hypertension), adding the very early LC degeneration as a hypothetical starting point of protein accumulation by impairing circadian BP regulation.

The present study presents some limitations. The main one is the relatively small number of participants recruited, the cross-sectional nature of the study, and the lack of a control group without cognitive impairment. However, participants were well-phenotyped with AD biomarker status available. Regarding neuroimaging methods, we acknowledge that although DTI-ALPS index has been widely used as a measure of glymphatic function, and it correlates with intrathecal administration of gadolinium (the gold standard method of glymphatic measure), it is an indirect measurement and might be less sensitive than such invasive methods. Another limitation is that the sample was heterogeneous in vascular risk factors and presence of hypertension. In addition to the standard methods to define dipping and non-dipping patterns with the commonly used SBP percentage of dip (<10%), we also constructed the normal/ one abnormal / two abnormal BP groups combining SBP and DBP patterns. Although this approach is non-standard, and we acknowledge that SBP and DBP are highly correlated, we performed secondary analyses using these groups to account for any deviation of sleep vascular BP given the exploratory nature of the study. On the other side, this study presents several strengths. It is the first one to address the association between sleep BP patterns and MRI glymphatic proxies and it was conceptualized within a wider theory of the interplay among glymphatic system and its vascular driver. Although some associations are observed, further longitudinal studies in larger samples are needed to confirm these findings.

In conclusion, our findings suggest that deviations of the physiological dipping sleep BP pattern in cognitively impaired individuals are associated to lower DTI-ALPS measures, a proxy of glymphatic dysfunction. Abnormal BP patterns also relate to presence of AD pathological biomarkers. These results imply that nighttime BP alterations may impact the functioning of glymphatic system, possibly contributing to pathological processes in AD. Our study opens new lines of research on therapeutic strategies targeting vascular health, such as antihypertensive treatments promoting physiological BP dip during sleep, that could have significant implications for slowing or preventing progression of neurodegenerative diseases.

## Acknowledgments

We would like to acknowledge the patients and families for their generous collaboration.

## Author contribution

Mariateresa Buongiorno: conception, data collection, analyses, and writing,

Gonzalo Sánchez-Benavides: analyses and writing

Giovanni Caruana: data collection, analyses, and critical review

Andrea Elias-Mas: data collection, analyses, and critical review

Cristina Artero: data collection

Natalia Cullell: analyses and critical review

Pilar Delgado: analyses and critical review

Darly Milena Giraldo: analyses and critical review

Clara Marzal-Espí: data collection and critical review

Oriol Grau-Rivera: analyses and critical review

Alejandro de la Sierra: analyses and critical review

Ariane Delgado-Sanchez: analyses and critical review

Nicola J.Ray: analyses and critical review

Jerzy Krupinski: data collection, analyses, and critical review

## Statements and Declarations

### Ethical considerations

The study was approved by the Ethical Committee from the Hospital Universitari MútuaTerrassa, Terrassa (Barcelona), Spain and conducted according to the local regulations. Written informed consent was obtained from all the participants and/or legal guardians for the study.

### Data availability

Data will be available upon request to the corresponding author.

## References

1 Pickering TG. Blood Pressure During Normal Daily Activities, Sleep, and Exercise. JAMA 1982; 247: 992.

2 de la Sierra A. Ambulatory blood pressure monitoring. Current status and future perspectives. Med Clínica (English Ed 2024; 163: 25–31.

3 Salles GF, Reboldi G, Fagard RH, Cardoso CRL, Pierdomenico SD, Verdecchia P et al. Prognostic Effect of the Nocturnal Blood Pressure Fall in Hypertensive Patients. Hypertension 2016; 67: 693–700.

4 Lempiäinen PA, Ylitalo A, Huikuri H, Kesäniemi YA, Ukkola OH. Non-dipping blood pressure pattern is associated with cardiovascular events in a 21-year follow-up study. J Hum Hypertens 2024; 38: 444–451.

5 Gavriilaki M, Anyfanti P, Mastrogiannis K, Gavriilaki E, Lazaridis A, Kimiskidis V et al. Association between ambulatory blood pressure monitoring patterns with cognitive function and risk of dementia: a systematic review and meta-analysis. Aging Clin Exp Res 2023; 35: 745–761.

6 Tan X, Sundström J, Lind L, Franzon K, Kilander L, Benedict C. Reverse Dipping of Systolic Blood Pressure Is Associated with Increased Dementia Risk in Older Men: A Longitudinal Study over 24 Years. Hypertension 2021; 77: 1383–1390.

7 Chen H-F, Chang-Quan H, You C, Wang Z-R, Hui W, Liu Q-X et al. The circadian rhythm of arterial blood pressure in Alzheimer disease (AD) patients without hypertension. Blood Press 2013; 22: 101–5.

8 Wang H, Xu Y, Ren R, Yao F, Chen M, Sheng Z et al. Ambulatory Blood Pressure Characteristics of Patients with Alzheimer’s Disease: A Multicenter Study from China. J Alzheimer’s Dis 2021; 83: 1333–1339.

9 Riba-Llena I, Nafría C, Filomena J, Tovar JL, Vinyoles E, Mundet X et al. High daytime and nighttime ambulatory pulse pressure predict poor cognitive function and mild cognitive impairment in hypertensive individuals. J Cereb Blood Flow Metab 2016; 36: 253–263.

10 Guo H, Tabara Y, Igase M, Yamamoto M, Ochi N, Kido T et al. Abnormal nocturnal blood pressure profile is associated with mild cognitive impairment in the elderly: the J-SHIPP study. Hypertens Res 2010; 33: 32–36.

11 Tarumi T, Harris TS, Hill C, German Z, Riley J, Turner M et al. Amyloid burden and sleep blood pressure in amnestic mild cognitive impairment. Neurology 2015; 85: 1922–1929.

12 Iliff JJ, Wang M, Zeppenfeld DM, Venkataraman A, Plog BA, Liao Y et al. Cerebral arterial pulsation drives paravascular CSF-interstitial fluid exchange in the murine brain. J Neurosci 2013; 33: 18190–18199.

13 Nedergaard M, Goldman SA. Glymphatic failure as a final common pathway to dementia. Science (80-) 2020; 370: 50–56.

14 Mestre H, Tithof J, Du T, Song W, Peng W, Sweeney AM et al. Flow of cerebrospinal fluid is driven by arterial pulsations and is reduced in hypertension. Nat Commun 2018; 9: 4878.

15 Ray NJ, Cullell N, Clark O, Delgado-Sanchez A, Caruana G, Elias-Mas A et al. Glymphatic system health in early Alzheimer’s disease and its relationship to sleep, cognition and CSF biomarkers. 2024. doi:10.1101/2024.10.14.618324.

16 Jenkinson M, Beckmann CF, Behrens TEJ, Woolrich MW, Smith SM. FSL. Neuroimage 2012; 62: 782–790.

17 Smith SM, Jenkinson M, Woolrich MW, Beckmann CF, Behrens TE, Johansen-Berg H et al. Advances in functional and structural MR image analysis and implementation as FSL. Neuroimage 2004; 23 Suppl 1: S208–19.

18 Andersson JLR, Sotiropoulos SN. An integrated approach to correction for off-resonance effects and subject movement in diffusion MR imaging. Neuroimage 2016; 125: 1063–1078.

19 Taoka T, Naganawa S. Glymphatic imaging using MRI. J Magn Reson Imaging 2020; 51: 11–24.

20 Álvarez I, Aguilar M, González JM, Ysamat M, Lorenzo-Bosquet C, Alonso A et al. Clinic-Based Validation of Cerebrospinal Fluid Biomarkers with Florbetapir PET for Diagnosis of Dementia. J Alzheimer’s Dis 2018; 61: 135–143.

21 Rasmussen MK, Mestre H, Nedergaard M. Fluid transport in the brain. Physiol Rev 2022; 102: 1025–1151.

22 Mortensen KN, Sanggaard S, Mestre H, Lee H, Kostrikov S, Xavier ALR et al. Impaired Glymphatic Transport in Spontaneously Hypertensive Rats. J Neurosci 2019; 39: 6365–6377.

23 Xie L, Kang H, Xu Q, Chen MJ, Liao Y, Thiyagarajan M et al. Sleep drives metabolite clearance from the adult brain. Science 2013; 342: 373–377.

24 Fultz NE, Bonmassar G, Setsompop K, Stickgold RA, Rosen BR, Polimeni JR et al. Coupled electrophysiological, hemodynamic, and cerebrospinal fluid oscillations in human sleep. Science (80-) 2019; 366: 628–631.

25 Pase MP, Beiser A, Himali JJ, Tsao C, Satizabal CL, Vasan RS et al. Aortic Stiffness and the Risk of Incident Mild Cognitive Impairment and Dementia. Stroke 2016; 47: 2256–2261.

26 Heffernan KS, Wilmoth JM, London AS. Estimated Pulse Wave Velocity Is Associated With a Higher Risk of Dementia in the Health and Retirement Study. Am J Hypertens 2024; 37: 909–915.

27 Hughes TM, Kuller LH, Barinas-Mitchell EJM, McDade EM, Klunk WE, Cohen AD et al. Arterial Stiffness and β-Amyloid Progression in Nondemented Elderly Adults. JAMA Neurol 2014; 71: 562.

28 Hughes TM, Wagenknecht LE, Craft S, Mintz A, Heiss G, Palta P et al. Arterial stiffness and dementia pathology. Neurology 2018; 90. doi:10.1212/WNL.0000000000005259.

29 Kinjo Y, Saji N, Murotani K, Sakima H, Takeda A, Sakurai T et al. Enlarged Perivascular Spaces Are Independently Associated with High Pulse Wave Velocity: A Cross-Sectional Study. J Alzheimer’s Dis 2024; 101: 627–636.

30 Tarumi T, Thomas BP, Wang C, Zhang L, Liu J, Turner M et al. Ambulatory pulse pressure, brain neuronal fiber integrity, and cerebral blood flow in older adults. J Cereb Blood Flow Metab 2019; 39: 926–936.

31 Yu JH, Kim REY, Park SY, Lee DY, Cho HJ, Kim NHNH et al. Night blood pressure variability, brain atrophy, and cognitive decline. Front Neurol 2022; 13: 963648.

32 Giorgi FS, Galgani A, Puglisi-Allegra S, Limanaqi F, Busceti CL, Fornai F. Locus Coeruleus and neurovascular unit: From its role in physiology to its potential role in Alzheimer’s disease pathogenesis. J Neurosci Res 2020; 98: 2406–2434.

33 Mitsubayashi H, Kawamura H, Hara K, Miyagawa M, Kanmasse K. Locus Coeruleus is important for the Circadian Rhythms of Blood Pressure and Behavior in Spontaneously Hypertensive Rats. Jpn Heart J 1995; 36: 518–518.

34 Mitsubayashi H. Destruction of the locus coeruleus produces ultradian rhythm in spontaneously hypertensive rat. Am J Hypertens 2000; 13: S166–S167.

35 Braak H, Thal DR, Ghebremedhin E, Del Tredici K. Stages of the Pathologic Process in Alzheimer Disease: Age Categories From 1 to 100 Years. J Neuropathol Exp Neurol 2011; 70: 960–969.

36 Madelung CF, Meder D, Fuglsang SA, Marques MM, Boer VO, Madsen KH et al. Locus Coeruleus Shows a Spatial Pattern of Structural Disintegration in Parkinson’s Disease. Mov Disord 2022; 37: 479–489.

37 De Pablo-Fernandez E, Tur C, Revesz T, Lees AJ, Holton JL, Warner TT. Association of Autonomic Dysfunction With Disease Progression and Survival in Parkinson Disease. JAMA Neurol 2017; 74: 970.

38 Buongiorno M, Marzal C, Fernandez M, Cullell N, de Mena L, Sánchez-Benavides G et al. Altered sleep and neurovascular dysfunction in alpha-synucleinopathies: the perfect storm for glymphatic failure. Front Aging Neurosci 2023; 15: 1–8.

